# Knowledge, Attitude and Practices Regarding Digital Well-Being Features and their Association with Screen Time and Addiction in Maharashtra, India

**DOI:** 10.1101/2023.02.21.23286252

**Authors:** Amey Ambike, Shirish Rao, Raghav Paranjape, Shilpa Adarkar

**Affiliations:** Seth G.S. Medical College and K.E.M. Hospital, Mumbai, Maharashtra, India; Department of Psychiatry, Seth G.S. Medical College and K.E.M. Hospital, Mumbai, Maharashtra, India

**Keywords:** Digital Service Providers, Digital Well-being Features, Screen Addiction

## Abstract

**Background & Objective:** Digital Service Providers have come up with certain ‘Digital Well-Being Features’ as a step towards tackling harmful effects of screen overuse on physical and mental health. However, the awareness and use of the same remains scant. Our objective was to assess the knowledge, attitudes and practices regarding Digital Well-being features in the adult population of Maharashtra, India and the associations and correlations of the practice of using these features with screen time and degree of screen addiction.

**Methods:** A cross-sectional online questionnaire-based study was conducted among 335 participants who were selected using quota sampling and were administered a Smartphone Addiction Scale and a self-designed questionnaire.

**Results:** Knowledge attitudes were good and total of 65.4% participants were digital wellbeing feature users. Correlation of digital hygiene score and digital wellbeing score was found neither with addiction nor with the average screen time.

**Interpretation & Conclusion:** Knowledge, attitude and practices regarding Digital Well-Being features were adequate among the urban population of Maharashtra. However, their use was not found to be associated with reduced screen time or a low screen addiction score. With further development and standardization, these features can be a useful tool for prevention of screen overuse and addiction.

## Introduction

As the COVID-19 pandemic spread across the globe, many countries, including India, had to impose stay-at-home advisories and impose social distancing protocols and lockdown restrictions. Students took to online teaching-learning models as schools shut down, and the movement bans resulted in a higher use of screens for entertainment and social interactions. These measures inadvertently increased the usage of varied screens, including not just computers & video gaming, but also television and mobile devices. (1) There is also a strong evidence that different types of screen use modify the association with mental health symptoms that users may suffer from, like depression, anxiety, conduct disorders, and attention problems in children and youth (2). This “potential” addiction has already shown to have a myriad of effects on physical and mental health of humans; with it being a significant risk factor to land up in obesity (3) and resulting in headaches, fatigue & sleep disturbances (4), as well as being associated with lack of concentration (4), anxiety and depression (5).

To tackle this problem, many Digital Service Providers (DSPs) have come up with acceptable solutions, which includes certain ‘Digital Well-being features’, to curb the amount of time spent with screen devices. ‘Digital Wellbeing by Android’ is a feature available in all phones made by Samsung, Xiaomi, Realme3 Pro, Vivo S1, Oppo phones in their latest models (6,7,8,9,10). ‘Zen Mode’ by OnePlus is a similar feature available in all phones created by OnePlus (11). Similar Features are also available on Apple devices (12). Many applications like Your Hour, Moment App, Lock My Phone etc. (13,14,15) also have similar features, but no single app has all the desired features. Apart from a few commonly used social media applications like Facebook, Instagram and YouTube which do have features to reduce usage and limit screen-time, there are minimal to no digital well-being features available in any other social media, gaming or OTT streaming application.

The problem still revolves around the effectiveness and accessibility of the already existing tools and more importantly, the awareness about these features in the targeted audience. Well-known features need to be scrutinised again to reassess their effectiveness and to address their shortcomings. Hence, this study was undertaken to judge the level of awareness regarding such Digital Wellbeing tools and their perceived efficacy.

Our primary objective was to assess the knowledge, attitudes and practices regarding Digital Well-being Features in the adult population of Maharashtra, India. We also assessed the associations and correlations of the practice of using Digital Well-being Features with screen time and degree of screen addiction. Our secondary objective was to assess the difference in Knowledge, attitudes and practices regarding Digital Well-being Features with respect to socio-demographic factors.

## Materials & Methods

A cross-sectional online questionnaire-based study was conducted between March-October 2021. Individuals who were above the age of 18, residents of Maharashtra, had regular access to smartphones, were capable of reading and comprehending questions, and were freely willing to participate were included in the study. All those who were suffering from/ had history of severe psychiatric disorders were excluded as their health conditions may render them incapable of participating in the study in an effective manner.

### Sample size and sampling technique

A sample size of 384 participants was calculated. Assuming maximum variability, which is equal to 50% (p=0.5), 95% confidence level with ± 5% precision, using Cochran’s formula;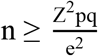 was used, where Z is the selected critical value of desired confidence level, p is the estimated proportion of an attribute, q = 1 – p and e is the desired level of precision.

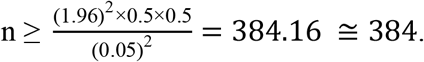

Participants were recruited by forwarding the link of online questionnaires via E-mail and social media platforms. Quota sampling was used and equal proportion of participants were recruited from age group 18-30 and more than 30 years.

### Ethical Considerations

The study commenced after the approval by the Institutional Ethics Committee (IEC) (EC/OA-170/2020). Identification details like name and address were not taken from the participant, keeping this an anonymous survey. Informed consent was obtained from the participants via the informed consent form attached before the questionnaire.

### Study Tools

#### Questionnaire

The questionnaires were developed with the aim of aiding comprehensive interpretation of thoughts and views of the participants of the survey and were available in English, Hindi and Marathi.The questionnaires included multiple choice questions, the options of which were carefully selected after analysing all possibilities such that the participants found their answers to the questions from the given options. It also included Yes/No type questions. It was divided into 3 parts: 1) Socio-Demographic information, 2) Knowledge, Attitudes & Perceptions 3) Practices.

#### Development & Validation questionnaire

The Questionnaire was structured by closely analysing the availability of Digital Well-being features in the Top mobile brands, social media, Gaming & OTT Platforms. Opinions of general public were considered by conducting a pilot study using an open-ended questionnaire. The content validation was done by experts from departments relevant to the topic of the study. The inputs and opinions given by them were thoroughly analysed and were incorporated into the questionnaires. The content validity ratio of the questionnaire was 0.967.

#### Smartphone Addiction Scale-Short Version (16)

Smartphone Addiction Scale - Short Version (SAS-SV) is a scale for smartphone addiction that consisted of 6 factors and 10 items with a six-point Likert scale (1: ‘‘strongly disagree’’ and 6: ‘‘strongly agree’’) based on self-reporting. The six factors were daily-life disturbance, positive anticipation, withdrawal, cyberspace-oriented relationship, overuse, and tolerance. The SAS-SV has showed good reliability and validity for the assessment of smartphone addiction. The internal consistency and concurrent validity of SAS-SV has been verified with a Cronbach’s alpha of 0. 911.The ROC analysis results showed an area under a curve (AUC) value of 0.963(0.888–1.000), a cut-off value of 31, sensitivity value of 0.867 and specificity value of 0.893 in males while an AUC value of 0.947(0.887–1.000), a cut-off value of 33, sensitivity value of 0.875, and a specificity value of 0.886 in females. High risk of smartphone addiction was classified at scores between 32-60 for males and 34-60 for females.

### Statistical Analysis

The data was obtained in the form of a spreadsheet, under Microsoft Excel 2019 and analysed using SPSS 26.0 software. The calculations were carried out in numbers and their percentages. The p values <0.05 were considered statistically significant. Unpaired t test was used for analysing the differences in the screen time & addiction scores between the two groups, who used & did not use the Digital Well-being Features. Pearson’s Correlation was used to find the correlation of screen time & addiction scores with the frequency of use of each Digital Well-being Feature. Chi-square test was used to assess the difference in Knowledge, Attitudes and Practices regarding Digital Well-being Features with respect to Socio-demographic factors.

## Results

Results were obtained from a total of 335 participants of whom 57% were males and 43% were females. The majority of the respondents (92%) were from urban areas, and only 8% respondents were from rural areas. 54.9% participants had an undergraduate degree, 34.3% had a postgraduate degree whereas rest were educated up to higher secondary level. Knowledge and Attitudes about various Digital Well-being Features and practice of digital hygiene have been summarized in the **Table 1**. App locking features (33%) followed by over-usage warnings (21%) were perceived to be the most effective to control screen usage by the participants **(Figure 1)**. 34.6% participants did not use any digital well-being features, 31.3% used in-built features in their phones, 6.9% used digital wellbeing applications whereas the remaining 27.2% used both in-built features and applications. Hence, a total of 65.4% participants (219) were Digital Well-being Feature users in our study. Practices regarding frequency of usage of Digital Well-being Features are summarized in **Table 2**. Blue light filter followed by time tracking were the most frequently used features whereas gray scale mode and Parental Control were least frequently used. No significant association of knowledge, attitudes or practices were found with socio-demographic factors (p>0.05).

**Table 1:** Knowledge and Attitudes about various digital well-being features and practice of Digital hygiene.

| Variables |  | Yes<br>n (%) | No<br>n (%) |
| --- | --- | --- | --- |
| **Knowledge** |  |  |  |
| **Digital Wellbeing Features** | Time tracking | 252 (75.2%) | 83 (24.8%) |
|  | Over usage warnings | 249 (74.3%) | 86 (25.7%) |
|  | App locking | 232 (69.3%) | 103 (30.7%) |
|  | Focus Mode | 186 (55.5%) | 149 (44.5%) |
|  | Phone locking | 204 (60.9%) | 131 (39.1%) |
|  | DND Mode | 290 (86.6%) | 45 (13.4%) |
|  | Blue light filter | 244 (72.8%) | 91 (27.2%) |
|  | Gray Scale Mode | 164 (49%) | 171 (51%) |
|  | Features in Facebook | 148 (44.2%) | 187 (55.8%) |
|  | Features in YouTube | 191 (57%) | 144 (43%) |
|  | Features in Instagram | 178 (53.1%) | 157 (46.9%) |
| **Attitudes** |  |  |  |
| **Digital Wellbeing Features help to** | Reduce aggregate screen time | 298 (89%) | 37 (11%) |
|  | Help focus on work and increase in productivity | 285 (85.1%) | 50 (14.9%) |
|  | Prevent Screen Addiction | 273 (81.5%) | 62 (18.5%) |
|  | Overcome Screen Addiction | 263 (78.5%) | 72 (21.5%) |
| **Suggestions for improving the digital wellbeing features** | Locking Features should be made stricter | 229 (68.4%) | 106 (31.6%) |
|  | Adding more Mental well-being features | 269 (80.3%) | 66 (19.7%) |
|  | Adding tools to monitor the level of screen dependency/addiction | 305 (91%) | 30 (9.0%) |
|  | Adding Helpline numbers/links to seek treatment | 206 (61.5%) | 129 (38.5%) |
|  | Creating a standardized app similar to Aarogya Setu for purpose of digital well being | 217 (64.8%) | 118 (35.2%) |
| **Practices** |  |  |  |
| **Digital Hygiene** | Avoiding devices upon waking up and before sleeping | 210 (62.7%) | 125 (37.3%) |
|  | Spending time with nature | 254 (75.8%) | 81 (24.2%) |
|  | Spending time with your family/ friends | 308 (91.9%) | 27 (8.1%) |
|  | Practicing a hobby/ sport | 271 (80.9%) | 64 (19.1%) |
|  | Practicing meditation | 195 (58.2%) | 140 (41.8%) |
|  | Self-care routines | 233 (69.6%) | 102 (30.4%) |
|  | Reading / Studying | 279 (83.3%) | 56 (16.7%) |
|  | Practicing Digital Detox | 145 (43.3%) | 190 (56.7%) |

**Table 2:**
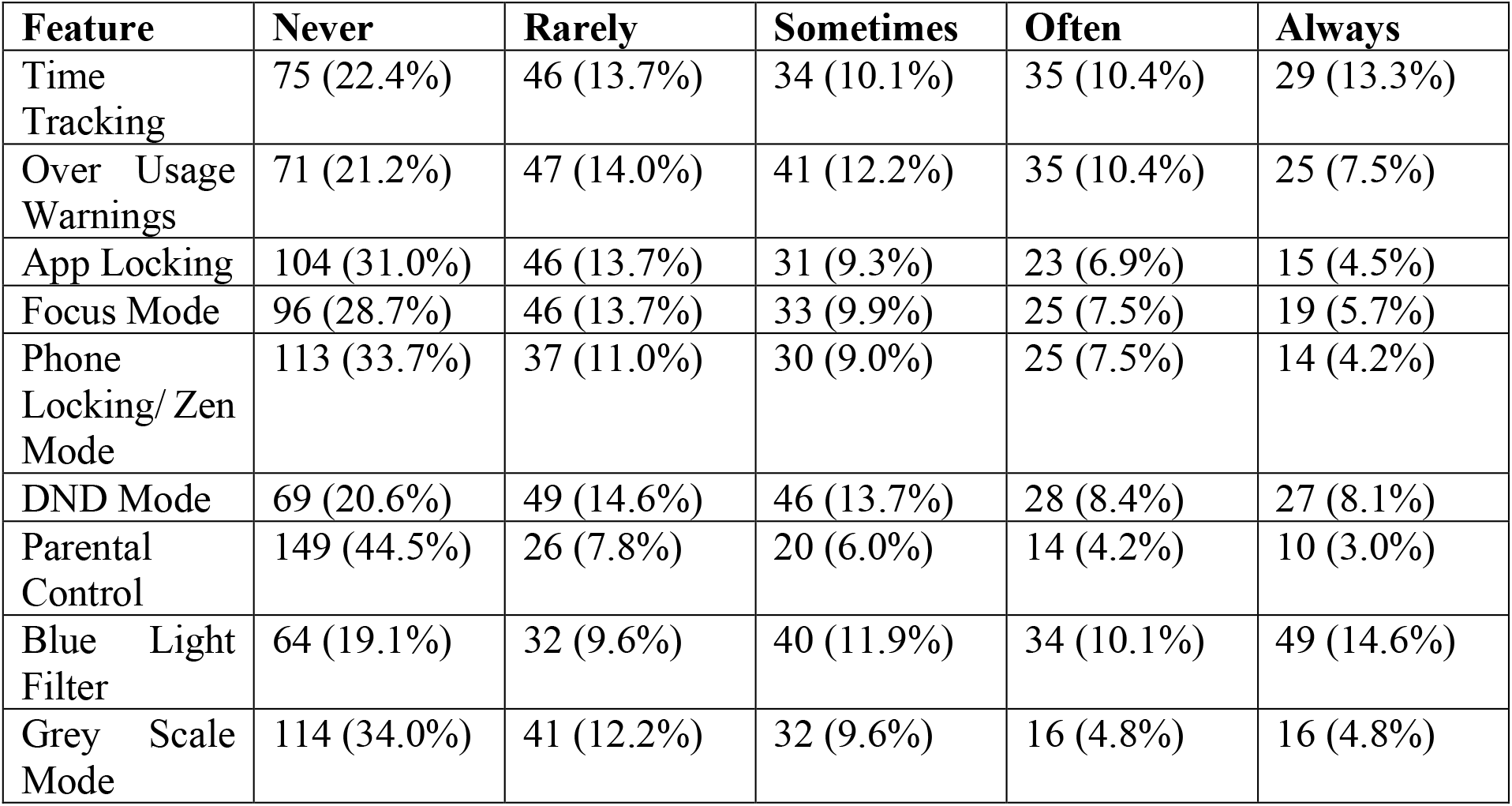
Frequency of using Digital well-being features.

**Figure 1:**
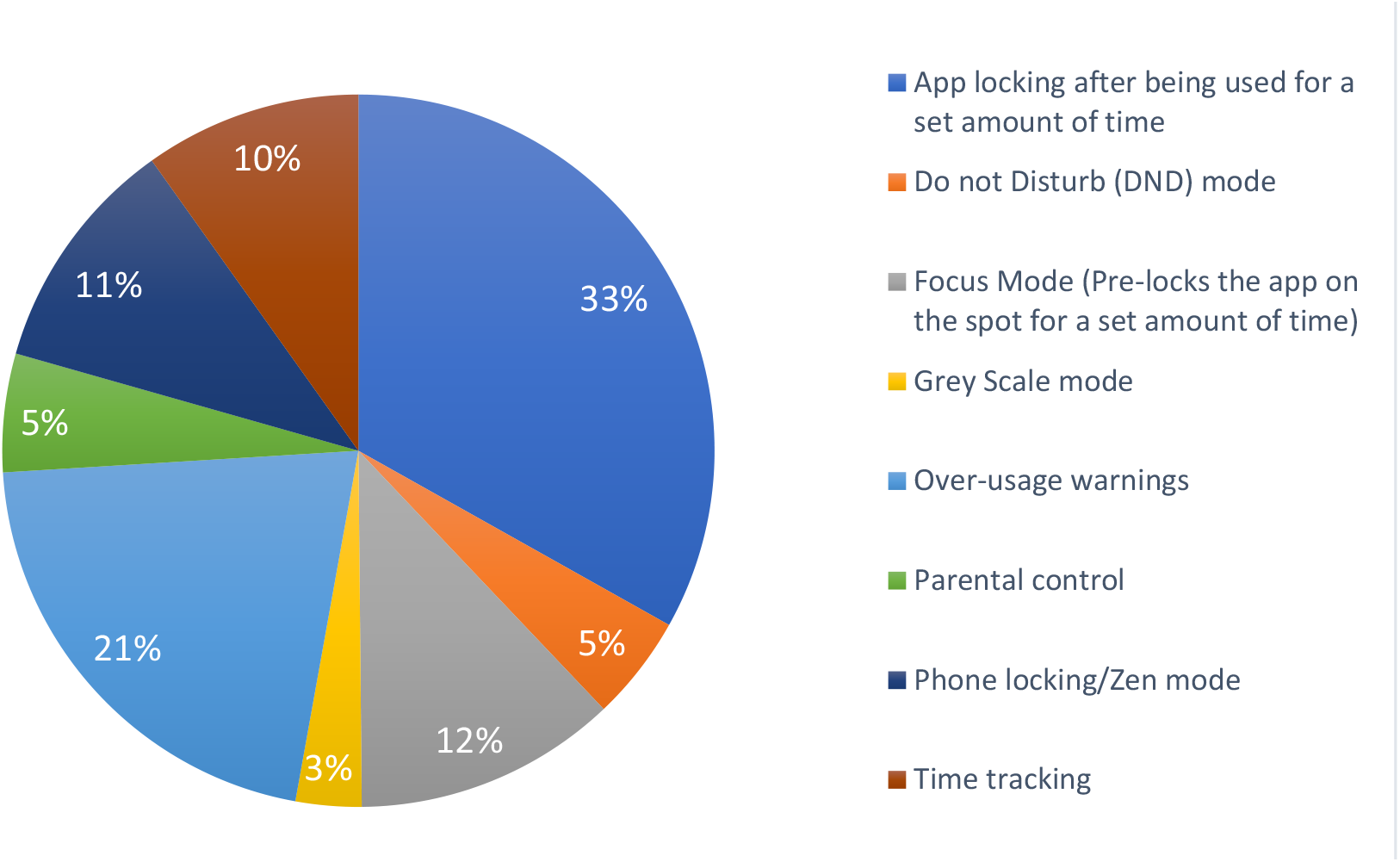
Digital well-being features perceived to be most effective to control screen usage.

When asked about adherence to app locking features, participants reported that, after exceeding the application usage limit, 28.1% participants stopped using the application, whereas 17.6% switched off the digital wellbeing features and continued using the application. On deciding to lock the phone but feeling the impulse to use it again, 29.3% kept the phone locked for the specified duration and only used it after the stipulated locking period ended, whereas 17.6% overrode the locking feature to use their phone again.

When the participants were screened for their smartphone addiction scores, 35% were addicts and rest were normal. No significant difference in their mean screen time (p=0.078) as well as addiction scores (p=0.112) were found between those who used and did not use digital wellbeing features **(Table 3)**. Though a weakly significant positive correlation (r=0.216, p=0.001) between addiction score and average screen time was found, correlation of digital hygiene score and digital wellbeing score was found neither with addiction scores (r= −0.84, p= 0.216; r= 0.014, p= 0.842) or the average screen time (r= 0.089. p= 0.192; r= −0.22, p=0.750) of the participants.

**Table 3:** Difference in mean screen time & addiction scores of Digital well-being feature users and non-users.

| Variable | Use of Digital Wellbeing Features | n (%) | Mean | Standard Deviation | t | P value | Mean Difference | 95% Confidence Interval of the Differences |  |
| --- | --- | --- | --- | --- | --- | --- | --- | --- | --- |
|  |  |  |  |  |  |  |  | Lower | Upper |
| Average Screen time per day in hours excluding work hours | Yes | 219 (65.3%) | 4.33 | 2.412 | 1.767 | 0.078 | 0.484 | -0.055 | 1.023 |
|  | No | 116 (34.7%) | 3.84 | 2.332 |  |  |  |  |  |
| Addiction Scores | Yes | 219 (65.3%) | 29.87 | 11.918 | 1.550 | 0.122 | 2.100 | -0.565 | 4.766 |
|  | No | 116 (34.7%) | 27.77 | 11.572 |  |  |  |  |  |

## Discussion

### Summary and Interpretation

More than two third of the participants were aware about Digital Well-being Features like time tracking, over usage warnings, app locking, phone locking, DND mode and blue light filter, indicating good knowledge among the study population. However, only half of the participants were also aware about the Focus mode, gray scale mode and inbuilt features in social media applications. An overwhelming majority believe that Digital Well-being Features can help reduce screen time, increase productivity and can be used to prevent as well as overcome screen addiction. Over 80% participants suggested that tools used to monitor level of screen addiction and mental well-being must be added to the currently available digital well-being features. Two thirds of the participants also suggested making the locking features stricter, addition of helplines and creation of a standard digital well-being smartphone application by the government. In the participant’s perception, app-locking features were voted to be most effective in regulating screen usage.

In terms of practices, about two thirds of the participants used at least one type of Digital Well-Being features. The features which had greater awareness among participants were used more frequently. These were Blue light filters, time tracking and over usage warnings. These features are less restrictive and require greater levels of self-control from the side of the user. Locking features being more restrictive were less frequently used. Among those who used locking features, about a fifth of them felt an uncontrollable urge to which they would act upon and find ways to unlock the apps/phone. Apart from features available on the device, the most common practices of digital hygiene adopted by the participants were spending screen-free time with family and peers or practicing a hobby or sports. Self-care routines and meditation even though highly beneficial for mental health, were least preferred to spend screen-free time.

When screened by the smartphone addiction scale, over a third of the participants were found to be at risk of addiction. However, neither their average screen time nor addiction score were associated with use of any of the Digital Well-being Features. The frequency of use of any of the Digital Well-being Features didn’t have a significant correlation with either their average screen time or addiction score. This indicates that even though participants perceived these features to be effective, statistically there is no difference in nature and duration of screen usage between digital well-being feature users and non-users. Interestingly participants with higher screen time had a higher frequency of using these features, indicating the presence of insight among the participants, but still an overall lack of impulse control.

### Implications

Currently, the presence of multiple non-standardized features may be reducing the usage frequency and thus effectiveness of these features. Standardizing these features and focusing on monitoring the screen addiction level over screen time would help participants differentiate between productive (work, study) usage and harmful usage. Considering good knowledge and attitudes among the population, digital well-being features at present in the form of time tracking, over usage warnings and locking features may best be used as a preventive strategy. The primary treatment modality for screen addiction is Cognitive-behavioral therapy (CBT) (17), but the problem remains about its awareness among the general public & their compliance with complete course of therapy. Inbuilt Digital well-being features could not only help individuals build healthy screen habits, but also the additional features like those of Smartphone Addiction Management System (SAMS) (18) could assist in preventing as well as treating screen addiction to a great extent. SAMS works by 1) Monitoring smartphone usage for 24 hours, without interruption, recording items including app use time, duration, location, and relevant information, such as a URL access location 2) Maintains records and uses a preprogramed analysis system to track the unhealthy or addictive screen routine & provide warnings 3) Provides Digital & mental well-being tips along with access to online therapy.

### Strengths and Limitations

Though many studies assessing the screen usage patterns, screen addiction and its health impacts have been conducted in India, this one of the first studies to assess the potential role of Digital Well-being Features available on the devices to prevent and treat screen addiction. The study has been conducted in an age-stratified representative sample from the state of Maharashtra. Being an initial study, it also has multiple limitations. The sample is mainly representative of adult urban population of the state of Maharashtra, hence the results can’t be generalized pan-India. Being an online study, the responses of the participants cannot be validated. There are chances of reporting as well as recall bias while entering the screen times, addiction scale scores and frequency of usage of well-being features. Being cross-sectional in nature, our study can neither assess the true frequency of usage of these features or the day-to-day screen time of the participant, thus it cannot conclude about the true efficacy of each of the Digital Well-being Features. Our participants also reported usage of different types of applications and in-built features for similar purposes which again makes it difficult to assess the efficacy of these features individually. Hence, our results can be used to design long-term large-scale cohort studies using standardized Digital Well-being Features, to assess their true efficacy and their potential in preventing and treating screen addiction.

## Conclusion

Knowledge, attitude and practices regarding Digital Well-Being Features were adequate among the urban population of Maharashtra. However, their use was not found to be associated with reduced screen time or low screen addiction score. With greater standardization of these features, they would be a useful tool for prevention of screen overuse and addiction. Results from our preliminary study can be used to design prospective studies to assess the true efficacy and utility of digital well-being features in prevention and treatment of screen addiction.

## Data Availability

All data produced in the present study are available upon reasonable request to the authors

## Acknowledgments

Nil

